# Automated Detection of Keratorefractive Laser Surgeries on Optical Coherence Tomography using Deep Learning

**DOI:** 10.1101/2024.03.08.24304001

**Authors:** Jad F. Assaf, Hady Yazbeck, Dan Z. Reinstein, Timothy Archer, Roland Assaf, Diego de Ortueta, Juan Arbelaez, Maria Clara Arbelaez, Shady T. Awwad

**Author notes:** Drs Jad F. Assaf and Hady Yazbeck contributed equally to this work. **Corresponding author:** Shady T. Awwad, MD Address: Department of Ophthalmology, American University of Beirut Medical Center Clinics, Beirut, Lebanon.

## Abstract

**PURPOSE:** To report a deep learning neural network on anterior segment optical coherence tomography (AS-OCT) for automated detection of different keratorefractive laser surgeries— including Laser In-Situ Keratomileusis with femtosecond microkeratome (Femto-LASIK), LASIK with mechanical microkeratome, photorefractive keratectomy (PRK), keratorefractive lenticule extraction (KLEx), and non-operated eyes—while also distinguishing the targeted ametropias, such as myopic and hyperopic treatments, within these procedures.

**DESIGN:** Cross-sectional retrospective study.

**METHODS:** A total of 14,948 eye scans from 2,278 eyes of 1,166 subjects were used to develop a deep learning neural network algorithm with an 80/10/10 patient distribution for training, validation, and testing phases, respectively. The algorithm was evaluated for its accuracy, F1-scores, area under precision-recall curve (AUPRC), and area under receiver operating characteristic curve (AUROC).

**RESULTS:** On the test dataset, the neural network was able to detect the different surgical classes with an accuracy of 96%, a weighted-average F1-score of 96% and a macro-average F1-score of 96%. The neural network was further able to detect hyperopic and myopic subclasses within each surgical class, with an accuracy of 90%, weighted-average F1 score of 90%, and macro-average F1-score of 83%.

**CONCLUSIONS:** Determining a patient’s keratorefractive laser history is vital for customizing treatments, performing precise intraocular lens (IOL) calculations, and enhancing ectasia risk assessments, especially when electronic health records are incomplete or unavailable. Neural networks can be used to accurately classify keratorefractive laser history from AS-OCT scans, a step in transforming the AS-OCT from a diagnostic to a screening tool in the refractive clinic.

## INTRODUCTION

Optical coherence tomography (OCT), a noncontact imaging technique, has revolutionized the visualization of biological tissues in vivo (1). Its ability to generate detailed cross-sectional images with quasi-histological resolution has been pivotal in the noninvasive clinical assessment of ocular structures, notably the cornea and anterior eye segment (2). The evolution of OCT, specifically anterior segment OCT (AS-OCT), aligns closely with the growing prevalence of corneal refractive surgeries. AS-OCT has emerged as a crucial tool in clinical practice, offering unparalleled accuracy in pre-operative diagnostics, surgical planning, and enhanced intra-operative imaging. It also plays a significant role in the post-operative evaluation and disease management (2), exemplified by its capacity to visualize the laser-assisted in situ keratomileusis (LASIK) flap during the early postoperative period (1).

In parallel, the field of artificial intelligence (AI), particularly deep learning, has seen a remarkable integration into AS-OCT applications. This integration marks a notable departure from the traditional focus on retinal OCT, expanding the scope of AI in ocular diagnostics. These advancements have proven effective in a range of clinical applications, from automated measurements such as ICL vault estimation (3,4), to sophisticated disease detection (5–7), and even in the creation of synthetic yet realistic corneal OCT images using deep learning (8).

A crucial aspect of clinical ophthalmology involves the accurate determination of a patient’s surgical history, particularly in refractive surgery. This information is essential not only for tailoring subsequent treatments but also for precise intraocular lens (IOL) calculations in cataract surgery and informing ectasia risk assessment algorithms (9,10).

The identification of a patient’s keratorefractive laser surgical history becomes paramount in cases where electronic health records are unavailable or incomplete. OCT B-scans provide a wealth of information in this regard. For instance, in LASIK patients, the presence of a flap in the anterior cornea, with distinct characteristics based on the cutting technique— femtosecond or microkeratome—can be identified. Mechanical keratomes usually produce meniscal flaps with deeper peripheral penetration and a more variable flap thickness (11,12), whereas femtosecond keratomes create flaps with uniform square peripheral edges with consistent and more predictable thickness across the cornea (13–16). In contrast, keratorefractive lenticule extraction (KLEx) surgeries exhibit a cap interface, without peripheral corneal surface penetration except at the small side-cut incision, differing from the LASIK hinge (17). In parallel, photorefractive keratectomy (PRK) and Phototherapeutic Keratectomy (PTK) treatments are characterized by the absence of Bowman’s layer, without any flap or interface (18).

This manuscript introduces a deep-learning neural network tailored for AS-OCT, devised for the automated detection and classification of various keratorefractive laser surgeries. The aim was for the network to categorize OCT B-scans into broad surgical classes, including non-operated eyes, femto-LASIK, mechanical LASIK, PRK/PTK, and KLEx, and to further discriminate between myopic and hyperopic corrections within each surgery class.

## METHODS

### Study Design and Ethical Compliance

This retrospective study analyzed anonymized OCT data from four international centers: the American University of Beirut (AUBMC), Lebanon; London Vision Clinic, United Kingdom; Muscat Laser Eye Center, Oman; and Aurelios Augenzentrum, Germany. Ethical approval was granted by the Institutional Review Board at AUBMC (IRB ID: BIO-2022-0038). Exemption from IRB was granted at the London Vision Clinic and Aurelios Augenzentrum. At Muscat Eye Laser Center, informed consent was obtained, as per routine, for using anonymized clinical data for research. Our study conforms to the principles of the Declaration of Helsinki and written informed consent was obtained from all patients prior to any procedure.

### Patient Selection and Data Preparation

The study encompassed preoperative and postoperative AS-OCT scan data from patients who underwent various types of keratorefractive laser surgeries. Eligibility for inclusion required that AS-OCT scans be conducted utilizing the MS-39 platform (CSO, Florence, Italy) and that participants had undergone one of the following keratorefractive surgeries: Femto-LASIK, LASIK with mechanical microkeratome, PRK/PTK, or KLEx, as well as those classified as non-operated eyes for normal control. KLEx surgeries included corneal lenticule extraction for advanced refractive correction (CLEAR, Ziemer) and small-incision lenticule extraction (SMILE, Carl Zeiss Meditec AG). Additionally, the type of ablation (myopic or hyperopic) was determined by the spherical equivalent correction (SE) value. Exclusion criteria included patients with multiple laser surgeries in the same eye, significant image artifacts, or minor refractive corrections (SE less than 1 diopter) since their changes on OCT might be too subtle to detect.

### Data Allocation and Preprocessing

Data were allocated to training, validation, or testing sets, ensuring no patient overlap across sets to prevent data leakage (19). The division ratio of patients was 80% training, 10% validation, and 10% testing. Radial B-scan images from varying angles were collected. Minority classes were oversampled for balanced representation. Image preprocessing included cropping to a 10mm corneal section, resizing to 512 x 512 pixels, and normalization using the training dataset’s color channel statistics. Data augmentation involved random alterations in rotation, contrast, and brightness.

### Deep Learning Model Architecture and Training

A convolutional neural network utilizing the ResNet18 architecture was employed (20). The model was pre-trained on ImageNet, a comprehensive image database, to facilitate high-level image feature detection (21). Transfer learning from a pre-trained model expedites training and reduces data requirements. The ResNet18 convoluted neural network (CNN) connected to a 512-neuron dense layer with 40% dropout regularization and an 8-neuron output layer, correlating to the various surgical classes. The model had 11, 710, 024 trainable parameters. Training involved 100 epochs, weighted cross-entropy loss, AdamW optimization, and a batch size of 32. Patient class ratios weighted the loss function.

### Optimization and Implementation

Optimal learning rates were determined through a range test (22), with discriminative fine-tuning applied to the ResNet18 layers (23). The ‘1cycle’ learning rate scheduler was used for efficiency (24). The model was implemented in Python using PyTorch (version 1.13.1+cu117) (25) and parallelized on 2 RTX 3080 GPUs (NVIDIA, USA), donated by Hugging Face (New York, USA).

### Evaluation and Statistical Analysis

The model’s performance was evaluated against the validation set after each epoch to monitor for overfitting. The model at the iteration with minimal validation loss was further assessed on the independent test set.

Comprehensive statistical evaluations were facilitated by the TorchMetrics package (version 0.9.3), incorporating a suite of metrics. Among these, receiver operating characteristic (ROC) curves plot the true positive rate against the false positive rate at varied threshold settings, elucidating the model’s discriminative capacity between classes. Precision-recall (PR) curves, mapping the precision (the ratio of true positive results to all positive predictions) against recall (the ratio of true positive results found in all relevant instances), are particularly insightful for models trained on imbalanced datasets.

The F1 score, harmonizing precision and recall, was computed in two variations: macro and weighted. The macro F1 score averages the F1 scores of each class, treating all classes equally regardless of their sample size. Conversely, the weighted F1 score accounts for class imbalance by weighting each class’s F1 score by its presence in the dataset, offering a measure that reflects the model’s performance across the unevenly distributed classes.

Further, we calculated both macro and weighted one-vs-one AUC scores to evaluate the model’s performance. The macro-AUC score averages the area under the ROC curve for each class, disregarding class imbalance, thus providing a generalized metric of the model’s ability to classify each class against the rest. The weighted one-vs-one AUC score, on the other hand, computes the AUC for each pairwise class comparison, weighting the contribution of each class pair by the prevalence of the respective classes in the dataset. This nuanced metric offers insight into the model’s classification prowess, particularly in distinguishing between similar classes in an imbalanced dataset.

Additional metrics, such as overall accuracy and confusion matrices, rounded out the evaluation, providing a multidimensional view of the model’s predictive performance.

### Model Output Contextualization

In our analysis, we initially developed an 8-class model to meticulously classify keratorefractive surgeries by both procedure type and the ametropia addressed. The categories delineated were pre-operative, myopic Femto-LASIK, hyperopic Femto-LASIK, myopic mechanical LASIK, hyperopic mechanical LASIK, myopic PRK or PTK, hyperopic PRK or PTK, and KLEx. This detailed classification scheme allowed for a nuanced understanding of the model’s ability to distinguish among a broad spectrum of keratorefractive surgeries.

Subsequently, to streamline the model’s utility for broader applications, we condensed the 8-class model into a 5-class variant. This consolidation was achieved by merging categories based on the type of procedure while disregarding the specific ametropia treated. Hence, myopic and hyperopic Femto-LASIK were combined into a single Femto-LASIK category, similarly for mechanical LASIK, and PRK/PTK categories, leading to a simplified classification comprising pre-operative, Femto-LASIK, mechanical LASIK, PRK/PTK, and KLEx. This approach underscores that the model’s robustness in the detailed 8-class variant directly substantiates its validity for the more generalized 5-class model.

Given that the model’s decision-making process is based on individual B-scan images and considering that multiple radial B-scans are available per patient, a majority selection algorithm was incorporated for eye-level classification. This entails aggregating individual B-scan predictions in a voting mechanism to determine the predominant classification for each eye.

## RESULTS

A total of 14,948 OCT B-scans from 2,278 eyes of 1,166 patients were used for the analysis. Table 1 provides the number of B-scans, patients, and eyes for each of the surgical classes. The training set consisted of 12,109 OCT B-scans from 1812 eyes of 930 patients. The validation set consisted of 1452 scans of 233 eyes of 118 patients. Training was performed with a starting learning rate of 10^e-3^ and for a total of 40 minutes. The model with the lowest validation loss was selected and tested using the testing set of 1387 eye-scans from 233 eyes of 118 patients.

**Table 1:** Database patient distribution across the different surgical classes and ametropia treated.

| Class | Patients | Eyes | Images |
| --- | --- | --- | --- |
| Preop | 410 | 807 | 3174 |
| Femto-LASIK – Myopic | 170 | 329 | 1172 |
| Femto-LASIK – Hyperopic | 90 | 173 | 1256 |
| Mechanical LASIK – Myopic | 137 | 272 | 3551 |
| Mechanical LASIK – Hyperopic | 43 | 85 | 1150 |
| PRK/PTK – Myopic | 147 | 285 | 1422 |
| PRK/PTK – Hyperopic | 59 | 111 | 2433 |
| KLEx | 110 | 216 | 790 |
| **Total** | **1166** | **2278** | **14948** |

For the analysis of the testing set, the 5-way prediction model for surgical classes achieved an accuracy of 96%, a macro F1 average of 96% and a weighted F1 average of 96% (Fig. 1A and Table 2A). The 8-way prediction model for surgical classes and ametropia treated achieved an accuracy of 90%, a macro F1 average of 83%, a weighted F1 average of 90% (Fig. 1B and Table 2B). ROC curves and PR curves for the 8-way prediction model are shown in figures 2A and 2B, respectively. The one-vs-one ROC AUC scores were 97.18% (macro) and 97.89% (weighted), and one-vs-rest ROC AUC scores were 97.64% (macro) and 98.63% (weighted).

**Figure 1A:**
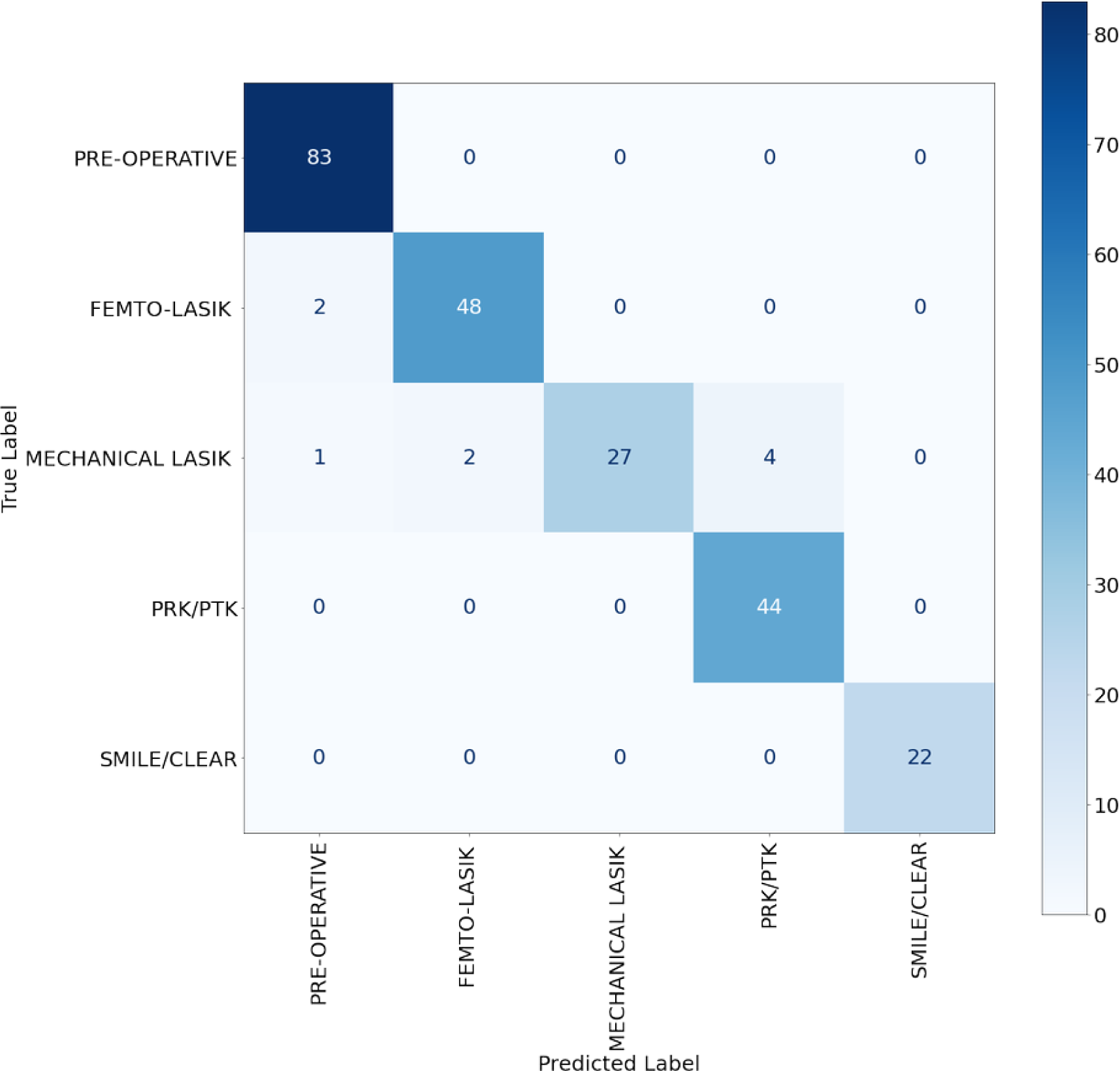
5-way classification confusion matrix on the test set.

**Figure 1B:**
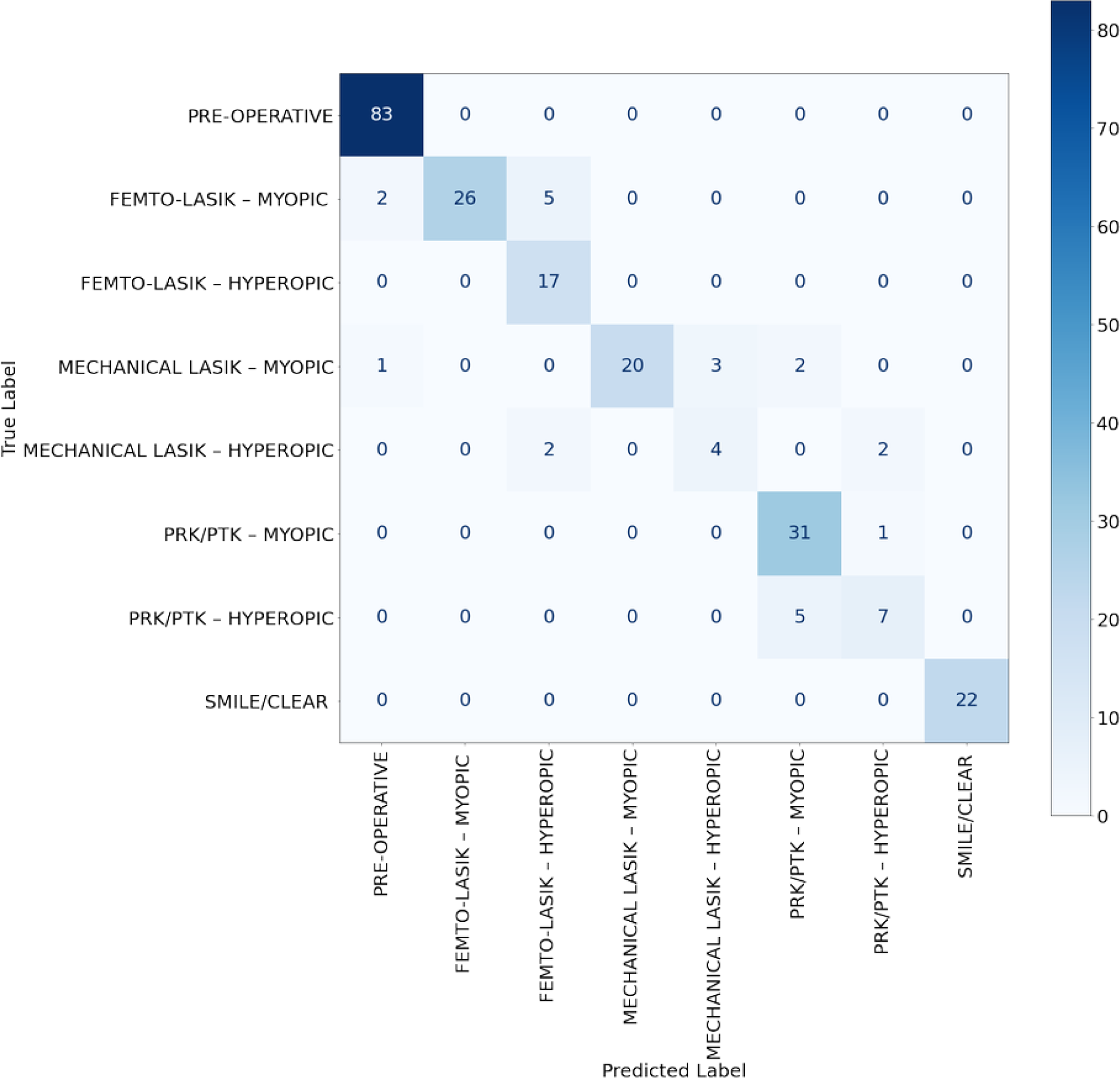
8-way classification confusion matrix on the test set.

**Figure 2A:**
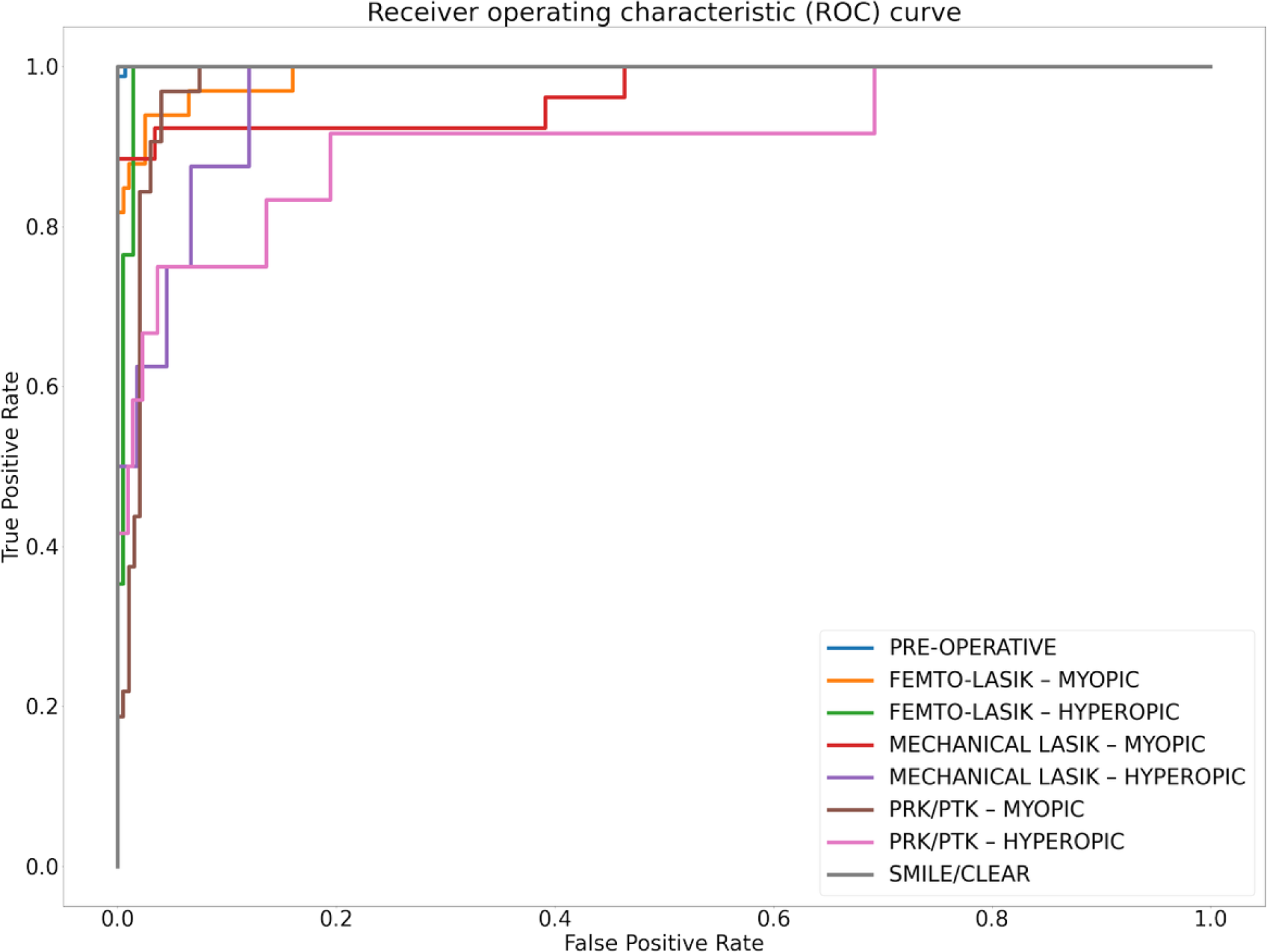
8-way ROC on the test set.

**Figure 2B:**
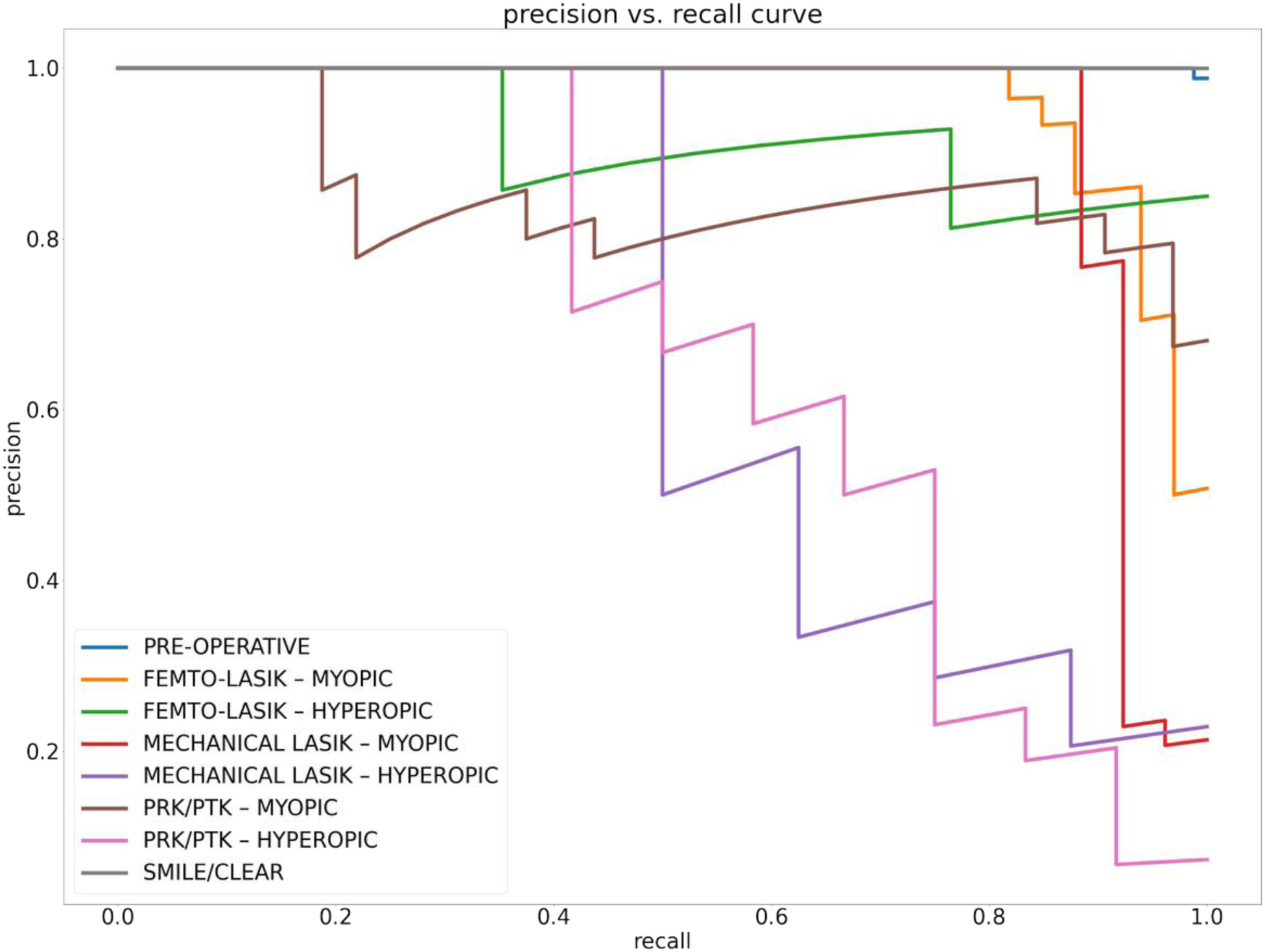
8-way PR curve on the test set.

**Table 2A:** 5-way classification results on the test set.

|  | PRECISION | RECALL | F1-SCORE | NUMBER OF EYES |
| --- | --- | --- | --- | --- |
| PRE-OPERATIVE | 0.97 | 1.00 | 0.98 | 83 |
| FEMTO-LASIK | 0.96 | 0.96 | 0.96 | 50 |
| MECHANICAL LASIK | 1.00 | 0.79 | 0.89 | 34 |
| PRK/PTK | 0.92 | 1.00 | 0.96 | 44 |
| KLEx | 1.00 | 1.00 | 1.00 | 22 |
| ACCURACY |  |  | 0.96 | 233 |
| MACRO AVG | 0.97 | 0.95 | 0.96 |  |
| WEIGHTED AVG | 0.96 | 0.96 | 0.96 |  |

**Table 2B:** 8-way classification results on the test set.

|  | PRECISION | RECALL | F1-SCORE | NUMBER OF EYES |
| --- | --- | --- | --- | --- |
| PRE-OPERATIVE | 0.97 | 1.00 | 0.98 | 83 |
| FEMTO-LASIK – MYOPIC | 1.00 | 0.79 | 0.88 | 33 |
| FEMTO-LASIK – HYPEROPIC | 0.71 | 1.00 | 0.83 | 17 |
| MECHANICAL LASIK – MYOPIC | 1.00 | 0.77 | 0.87 | 26 |
| MECHANICAL LASIK – HYPEROPIC | 0.57 | 0.50 | 0.53 | 8 |
| PRK/PTK – MYOPIC | 0.82 | 0.97 | 0.89 | 32 |
| PRK/PTK – HYPEROPIC | 0.70 | 0.58 | 0.64 | 12 |
| KLEx | 1.00 | 1.00 | 1.00 | 22 |
| ACCURACY |  |  | 0.90 | 233 |
| MACRO AVG | 0.85 | 0.83 | 0.83 |  |
| WEIGHTED AVG | 0.91 | 0.90 | 0.90 |  |

## DISCUSSION

This study has successfully developed a deep learning neural network that demonstrates proficiency in identifying a spectrum of keratorefractive laser surgeries from OCT B-scans. By leveraging transfer learning, the model adeptly managed dataset imbalances, showcasing a robust ability to classify between operative and non-operative eyes and to distinguish among specific types of surgeries.

In the 5-way prediction model, the network achieved an impressive accuracy of 96%. Yet, LASIK surgeries performed with mechanical keratomes experienced slightly higher misclassification rates. This trend may reflect the dataset’s composition, wherein mechanical LASIK procedures generally precede those done with femtosecond technology, resulting in older, well-healed surgical flaps that pose detection challenges for the model. This scenario highlights the complexity of detecting certain procedures, especially older ones, where the passage of time may diminish the distinctiveness of surgical signatures on OCT scans.

In the 8-way prediction, the network displayed capability in discerning the correction type within each surgical class, although with slightly diminished accuracy (90%), especially for the hyperopic variations of mechanical LASIK and PRK. These specific surgeries, being less common in our dataset, underscore the challenges posed by class imbalance and data scarcity. Attempts to mitigate this through transfer learning and adjusted loss function weighting were somewhat effective. The detection challenges are further exacerbated in hyperopic PRK cases due to the peripheral nature of the ablation and reduced OCT signal at the cornea’s edges due to increased angle of incidence, complicating the identification of Bowman’s layer changes. In such cases, a hyperopic PRK B-scan will be very similar to a normal patient. However, the model did not confuse both classes and the main errors arose in distinguishing myopic and hyperopic variations among each surgery.

An essential aspect of refining our model’s accuracy involves understanding the subtle distinctions between different keratorefractive treatments, particularly when distinguishing between myopic and hyperopic corrections. This involves analyzing corneal epithelial thickness variations. Myopic ablations typically present with a compensatory thicker central epithelium (26), whereas hyperopic ablations exhibit thinner central epithelium due to induced corneal steepening (27,28). In PRK corrections, hyperopic treatments typically spare Bowman’s layer in the very center, while myopic PRK treatments ablate Bowman’s layer along all of the optical zone.

To illustrate these distinctions further, Figure 3 presents OCT B-scans of various keratorefractive procedures, applanated along the anterior corneal curvature, highlighting the range of surgeries addressed in this study. Applanation enhances the visualization of variations in epithelial thickness and provides tomographic views of flap and cap interfaces.

**Figure 3:**
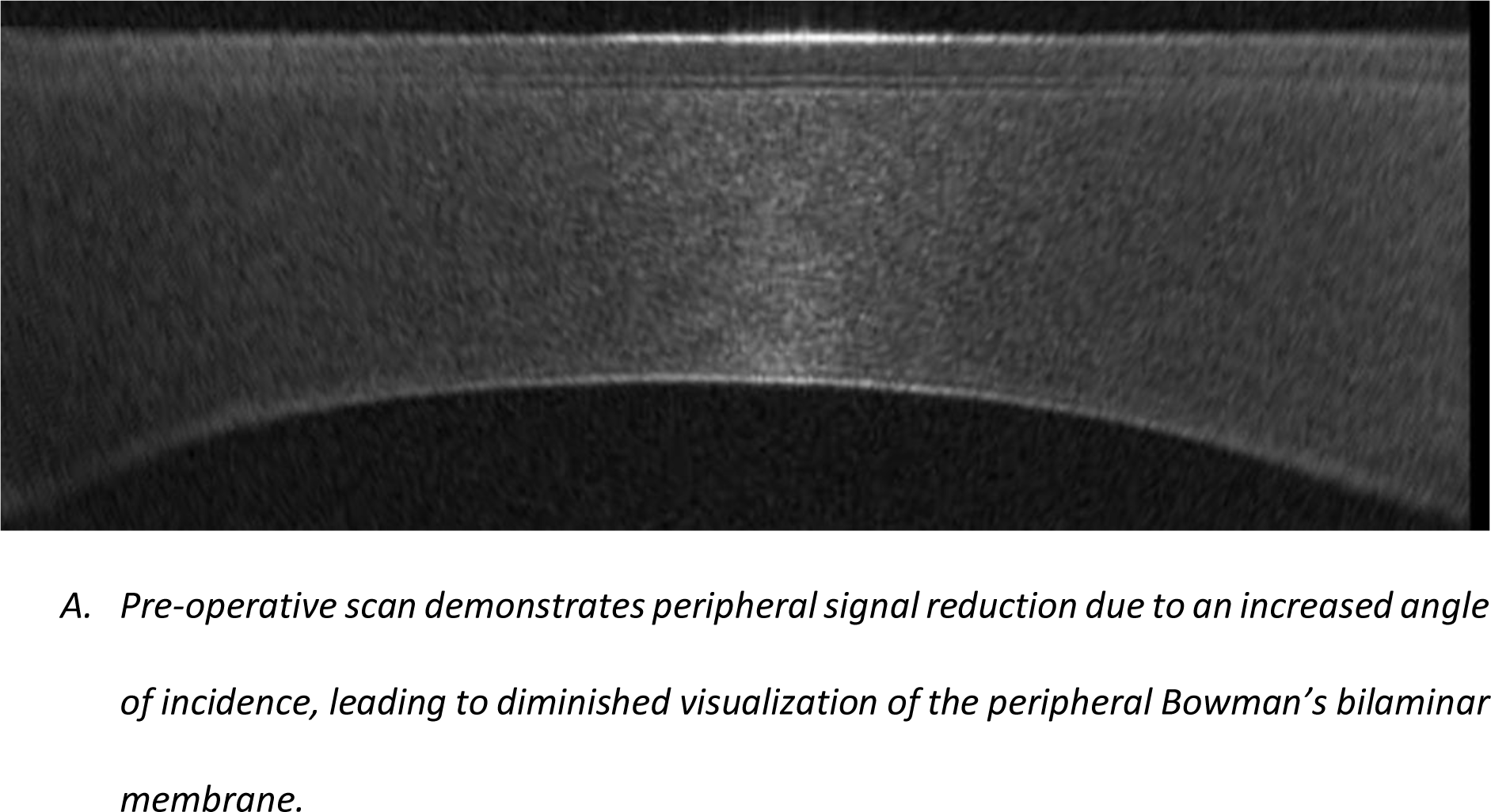
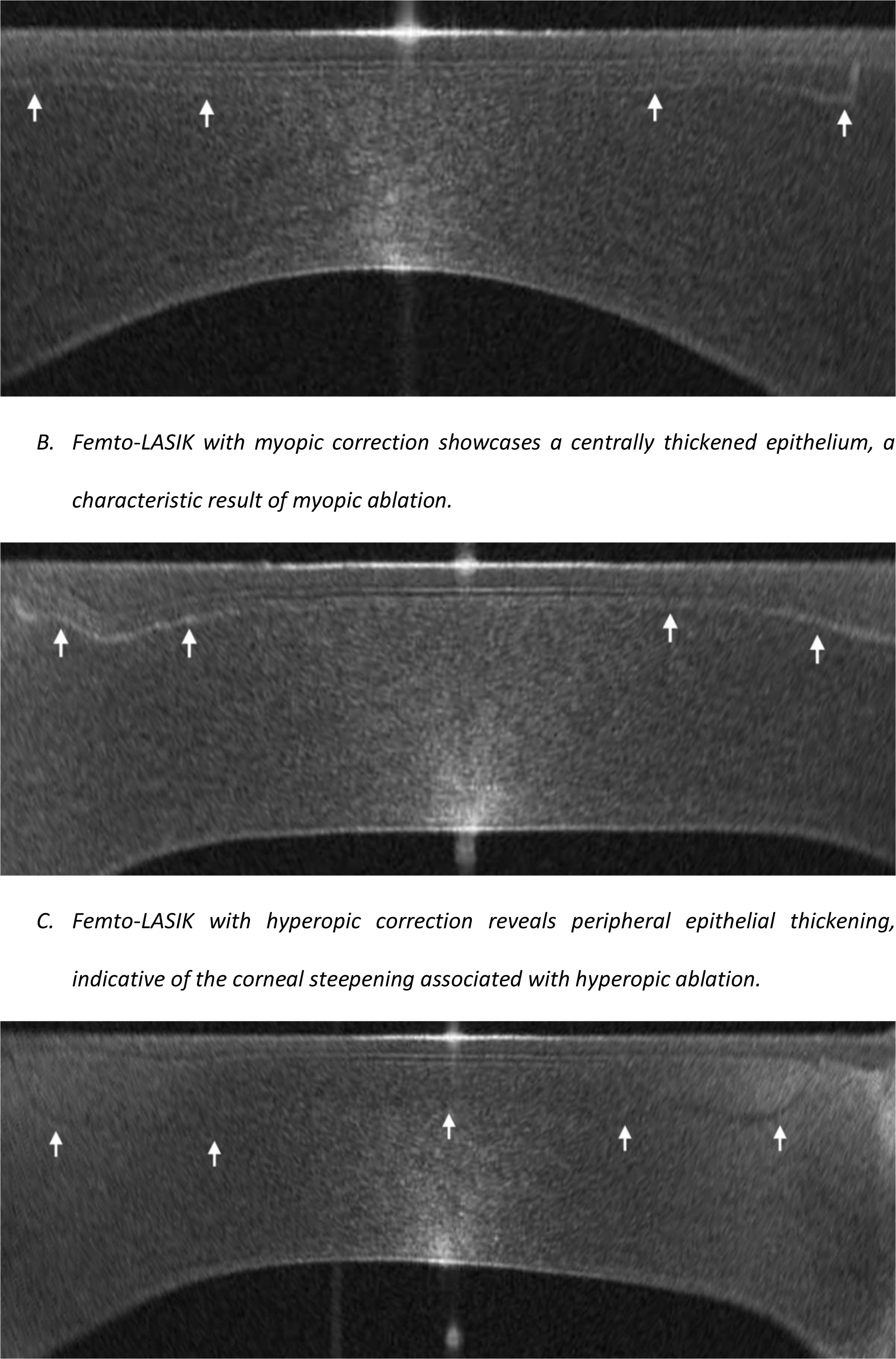
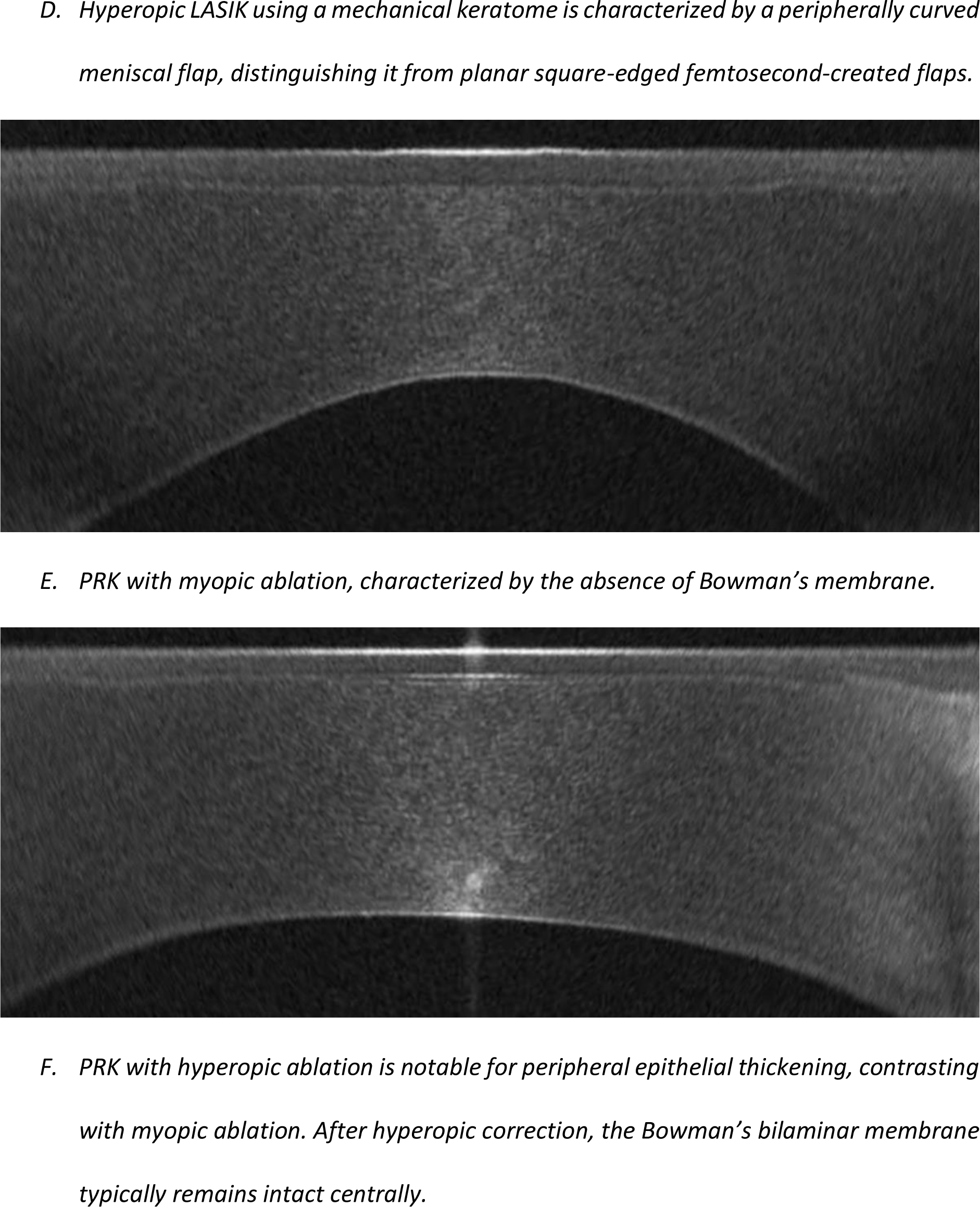
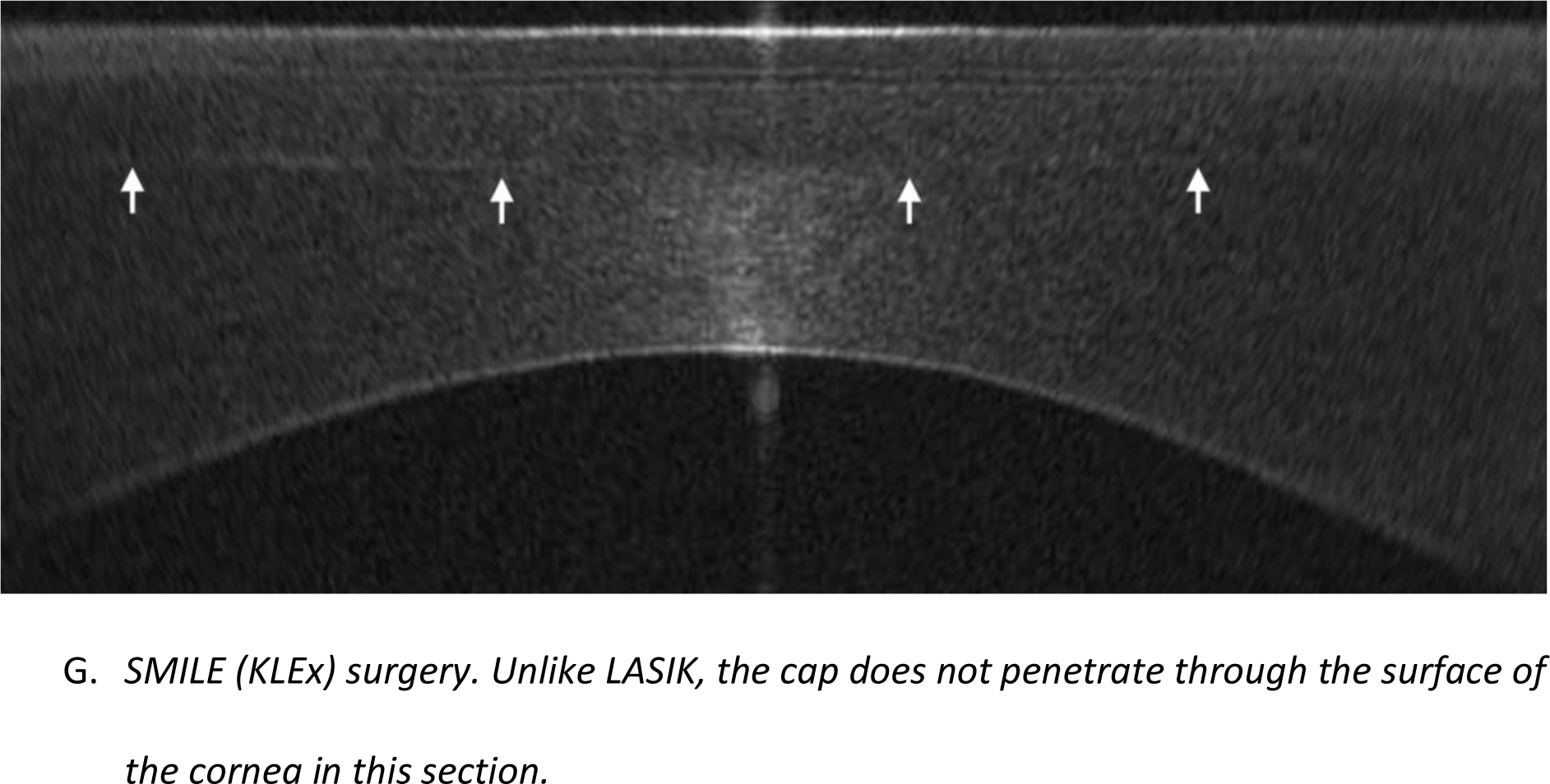
OCT B-scans depicting various keratorefractive procedures, applanated along the anterior corneal curvature. Arrows outline the flap/cap interfaces.

The sole reliance on B-scans is a notable limitation of this study. While radial B-scans yield a wealth of information, integrating tomographic parameters such as pachymetry, epithelial thickness, and corneal curvature could enrich the model’s understanding of the cornea’s refractive status. The majority voting mechanism used for classification does not consider spatial dependencies inherent in these scans, which could be addressed by a network architecture that integrates all B-scans collectively.

Enhancements to the data preprocessing approach, such as applanating the cornea, could sharpen our focus on variations in central and paracentral epithelial thickness—key indicators for distinguishing between myopic and hyperopic corrections (27,28). Furthermore, exploring the estimation of the extent of ablation performed on the cornea as well as the presence of a mixed astigmatism correction could serve as valuable features for future investigations. Furthermore, evolving our model to detect multiple sequential surgeries, including a primary LASIK followed by a PRK touch-up, or even enhancements via flap lift, also emerges as an important next step. Such developments would require a meticulously labeled dataset, encompassing cases with these specific surgical histories.

Despite the outlined limitations, the model demonstrates significant promise, with minimal errors observed in its classifications. Future work will aim to automate the segmentation of LASIK flaps and lenticule cap interfaces, enhancing the utility of this model in post-operative assessments.

In conclusion, this study presents a significant advancement in the application of deep learning in refractive surgery, specifically in the identification and classification of keratorefractive laser surgeries through OCT. Our model demonstrates robust performance and offers a promising foundation for further refinement and application. By addressing the outlined limitations and exploring the proposed enhancements, future iterations of this model have the potential to substantially improve post-operative assessments and contribute to more personalized patient care strategies.

## Data Availability

All data produced in the present study are available upon reasonable request to the authors.

## Financial interest

Dr Reinstein is a consultant for Carl Zeiss Meditec (Carl Zeiss Meditec AG, Jena, Germany). Dr Reinstein is also a consultant for CSO Italia (Florence, Italy) and has a proprietary interest in the Artemis technology (ArcScan Inc, Golden, Colorado) through patents administered by the Cornell Center for Technology Enterprise and Commercialization (CCTEC), Ithaca, New York. Dr Awwad is a consultant for Carl Zeiss Meditec (Carl Zeiss Meditec AG, Jena, Germany). Drs Assaf and Awwad have financial interest in NeuralVision – FZCO (Dubai, UAE). In addition to the previously disclosed interests, a full patent has been filed by Drs Assaf, Awwad, and Yazbeck pertaining to the methodologies and technologies discussed in this study. The remaining authors have no proprietary or financial interest in the materials presented herein.

## Funding

This research was partially funded by the Suhail Muasher Endowed Medical Student Research Award.

## Acronyms

OCT: Optical Coherence Tomography
AS-OCT: Anterior Segment Optical Coherence Tomography
LASIK: Laser-Assisted In Situ Keratomileusis
Femto-LASIK: Femtosecond Laser-Assisted In Situ Keratomileusis
PRK: Photorefractive Keratectomy
KLEx: Keratorefractive Lenticule Extraction
CLEAR: Corneal Lenticule Extraction for Advanced Refractive Correction
SMILE: Small Incision Lenticule Extraction
IOL: Intraocular Lens
AI: Artificial Intelligence
AUBMC: American University of Beirut Medical Center
IRB: Institutional Review Board
SE: Spherical Equivalent
CNN: Convolutional Neural Network
ROC: Receiver Operating Characteristic
PR: Precision-Recall
AUC: Area Under the Curve

## CRediT Author Statement

- Jad F. Assaf: Investigation, Methodology, Software, Writing - Original Draft, Funding acquisition, Project administration.
- Hady Yazbeck: Investigation, Visualization, Data Curation, Writing - Original Draft.
- Dan Z. Reinstein: Resources, Writing - Review & Editing.
- Timothy Archer: Resources, Writing - Original Draft.
- Roland Assaf: Data Curation, Writing - Review & Editing.
- Diego de Ortueta: Resources, Writing - Review & Editing.
- Juan Arbelaez: Resources, Writing - Review & Editing.
- Maria Clara Arbelaez: Resources, Writing - Review & Editing.
- Shady T. Awwad: Conceptualization, Supervision, Writing - Review & Editing.

## Notes

### Author Declarations

IRB of the American University of Beirut gave ethical approval for this work. IRB of London Vision Clinic and Aurelios Augenzentrum waived ethical approval for this work.

